# Cost-effectiveness of RSVpreF vaccine and nirsevimab for the prevention of respiratory syncytial virus disease in Canadian infants

**DOI:** 10.1101/2024.03.21.24304675

**Authors:** Gebremedhin B. Gebretekle, Man Wah Yeung, Raphael Ximenes, Alexandra Cernat, Alison E. Simmons, April Killikelly, Winnie Siu, Ellen Rafferty, Nicholas Brousseau, Matthew Tunis, Ashleigh R. Tuite

**Affiliations:** Centre for Immunization Programs, Public Health Agency of Canada, Ottawa, ON; Health Policy PhD Program, Faculty of Health Sciences, McMaster University, Hamilton, ON; Dalla Lana School of Public Health, University of Toronto, Toronto, ON; Institute of Health Economics, Edmonton, AB; Department of Medicine, Faculty of Medicine and Dentistry, University of Alberta, Edmonton, AB; Biological Risks Unit, Institut national de santé publique du Québec, Québec, Canada; Department of Social and Preventive Medicine, Université Laval, Québec, Canada

## Abstract

**Background:** Health Canada recently authorized the RSVpreF pregnancy vaccine and nirsevimab to protect infants against respiratory syncytial virus (RSV) disease.

**Objective:** Assess the cost-effectiveness of RSVpreF and nirsevimab programs in preventing RSV disease in infants, compared to a palivizumab program.

**Methods:** We used a static cohort model of a Canadian birth cohort during their first RSV season to estimate sequential incremental cost-effectiveness ratios (ICERs) in 2023 Canadian dollars per quality-adjusted life year (QALY) for nine strategies implemented over a one-year time period, from the health system and societal perspectives. Sensitivity and scenario analyses were conducted to explore the impact of uncertainties on the results.

**Results:** All-infants nirsevimab programs averted more RSV-related outcomes than year-round RSVpreF programs, with the most RSV cases averted in seasonal nirsevimab programs. Assuming list prices for these immunizing agents, all-infants nirsevimab and year-round RSVpreF programs were never cost-effective, with ICERs far exceeding commonly used cost-effectiveness thresholds. Seasonal nirsevimab with catch-up was cost-effective if prioritized for infants at moderate/high-risk (ICER <$28,000 per QALY) or those living in settings with higher RSV burden and healthcare costs (ICER of $5,700 per QALY). Using a $50,000 per QALY threshold, an all-infants nirsevimab program could be optimal if nirsevimab is priced at <$110-190 per dose. A year-round RSVpreF for all pregnant women/pregnant people plus nirsevimab for infants at high-risk was optimal if nirsevimab is priced at >$110-190 and RSVpreF priced at <$60-125.

**Interpretation:** Prophylactic interventions can substantially reduce RSV disease in infants, and targeted nirsevimab programs are the most cost-effective option at current product prices.

## INTRODUCTION

Respiratory syncytial virus (RSV) infection represents a substantial health and economic burden in infants and young children aged under five years globally, causing 3.6 million hospitalizations and 101,400 deaths annually (1). In Canada, 1% of all infants are hospitalized for RSV during the first year of life and RSV accounted for 9% of all hospital admissions in this population (2). Based on surveillance data from select Canadian pediatric hospitals, RSV causes over 2,500 hospital admissions annually, of which 50% occur within six months of age and 40% in the first two months (3). RSV is also estimated to cause up to 16 times more hospitalizations and emergency department visits in young children than influenza (4). Prematurity, age younger than one year, and underlying medical conditions (e.g., heart or lung disease) have been associated with elevated risk for severe RSV infection (5, 6).

The current standard of care for the prevention of RSV disease in Canada is palivizumab (SYNAGIS^TM^), a monoclonal antibody with a duration of protection of approximately one month, the use of which the National Advisory Committee on Immunization (NACI) recommended be restricted for infants and children at high risk for severe RSV disease entering their first or second RSV season (7). Palivizumab programs have limitations including high costs, complexity associated with the need for monthly dosing, and narrow eligibility criteria. Health Canada recently authorized two new products for the prevention of RSV lower respiratory tract infection (LRTI) in infants: nirsevimab (BEYFORTUS^TM^, Sanofi) and RSVpreF vaccine (ABRYSVO^TM^, Pfizer). Nirsevimab is a long-acting monoclonal antibody with a potential duration of protection of 5 months or longer that is directly administered to infants (8). RSVpreF is a prefusion F protein-based vaccine administered during pregnancy to protect newborns during the first few months after birth through passive transfer of transplacental antibodies (9). Another monoclonal antibody with a potential protection for several months, clesrovimab (MK-1654, Merck), is in a Phase clinical 3 trial expected to be completed in 2024 (10).

With the introduction of these new RSV prophylactic products, it is essential to determine the optimal strategy that provides the best value for money. In Canada, NACI publishes recommendations for vaccine programs, where economic evidence is one of several factors considered for decision-making (11). In this study, we conducted a model-based economic evaluation to evaluate the cost-effectiveness of multiple immunization strategies for protecting Canadian infants against RSV disease under various scenarios.

## METHODS

### Model overview

We conducted a model-based cost-utility analysis to assess the cost-effectiveness of using a single dose of RSVpreF vaccine in pregnant women and pregnant people and/or nirsevimab in infants for the prevention of RSV-associated outcomes in Canadian infants, compared to the current standard of care (i.e., palivizumab for infants at high-risk). A static cohort model was used to compare the health and economic impact of different RSV disease prevention strategies, from both the health system and societal perspectives, consistent with NACI guidelines (11). Outcome measures included a number of RSV-associated outcomes (i.e., outpatient healthcare provider visit, emergency department [ED] visit, pediatric ward hospitalization or intensive care unit [ICU] admission, death) averted, quality-adjusted life years (QALYs) lost, costs (in 2023 Canadian dollars), and incremental cost-effectiveness ratios (ICERs), defined as incremental cost per incremental health effect, expressed as dollars per quality-adjusted life-year (QALY) gained. For RSV-associated costs and outcomes that accrue over more than a year, we applied an annual discount rate of 1.5% (11). Since Canada does not have an explicit cost-effectiveness threshold (11), the study assessed cost-effectiveness of the interventions using commonly used thresholds of $50,000 and $100,000 per QALY. The study was designed, conducted, and reported following Canadian Guidelines for the economic evaluation of vaccination programs (11).

### Interventions, program scenarios, and timing of administration

We evaluated the impact of nine potential prevention strategies against RSV disease in infants entering their first RSV season. The model did not include infants entering their second RSV season. We modelled six nirsevimab programs, two RSVpreF vaccination programs and the palivizumab standard of care program:

i. Year-round nirsevimab program administered at birth for all infants
ii. Seasonal nirsevimab program without catch-up administered at birth for all infants born during the RSV season (i.e., from November to May)
iii. Seasonal nirsevimab program with catch-up in which all infants born during the RSV season receive their dose at birth and a catch-up dose is administered at the start of the RSV season (i.e., November) for all infants born outside of the RSV season (i.e., from June to October)
iv. Year-round nirsevimab program administered at birth for infants at moderate- and high-risk,
v. Seasonal nirsevimab program without catch-up administered at birth for infants at moderate- and high-risk born from November to May
vi. Seasonal nirsevimab program with catch-up in which infants at moderate- and high-risk born during the RSV season receive their dose at birth and a catch-up dose is administered at the start of the RSV season (i.e., November) for moderate- and high-risk born outside of the RSV season (i.e., from June to October)
vii. Year-round RSVpreF for all pregnant women and pregnant people
viii. Year-round RSVpreF for all pregnant women and pregnant people plus year-round nirsevimab offered to infants at high-risk (assuming no protection from RSVpreF)
ix. Current standard of care (i.e., palivizumab for infants at high-risk)

Prematurity is associated with an increased risk of RSV-related complications, and newborns were stratified into three groups based on their week of gestational age (wGA) at birth: infants at high-risk (extremely and very preterm, <33 wGA), infants at moderate-risk (late preterm, 33^0^ to 36^6^ wGA) and infants at low-risk (full term, ≥37^0^ wGA). Nirsevimab uptake among infants at moderate- and low-risk was assumed to be lower compared to infants at high-risk (70% vs. 80%, respectively) (12). RSVpreF was assumed to be administered year-round due to anticipated implementation challenges of a seasonal program. The timing of seasonal and year-round administration for the first season of RSV is provided in supplementary Figure 1. The national vaccination coverage rate for Tdap vaccine during pregnancy (64.8%) was used as an estimate of RSVpreF vaccine uptake (13). Palivizumab coverage was assumed to be the same as nirsevimab coverage among infants at high-risk (80%). Immunization coverage was assumed to remain constant across immunization calendar months.

### Model structure

The economic model structure consisted of three mutually exclusive health states: healthy, RSV-infected, and dead (Figure 1). The model followed monthly birth cohorts of Canadian newborns over a one-year time period and included lifetime costs and consequences for long-term outcomes (i.e., death due to RSV disease). Upon model entry, a proportion of the birth cohort was immunized based on each program’s estimated immunization coverage, assuming constant coverage across vaccination months. Infants could develop RSV disease over the model time horizon. Only medically-attended LRTI (MA-LRTI) was included in the model, with each episode resulting in either an outpatient healthcare provider visit, ED visit, hospitalization in pediatric general ward, or ICU. RSV-related death was assumed to occur only among infants who were hospitalized. The model was constructed and analyzed using TreeAge Software (TreeAge Software, Inc., Williamstown, MA).

**Figure 1.**
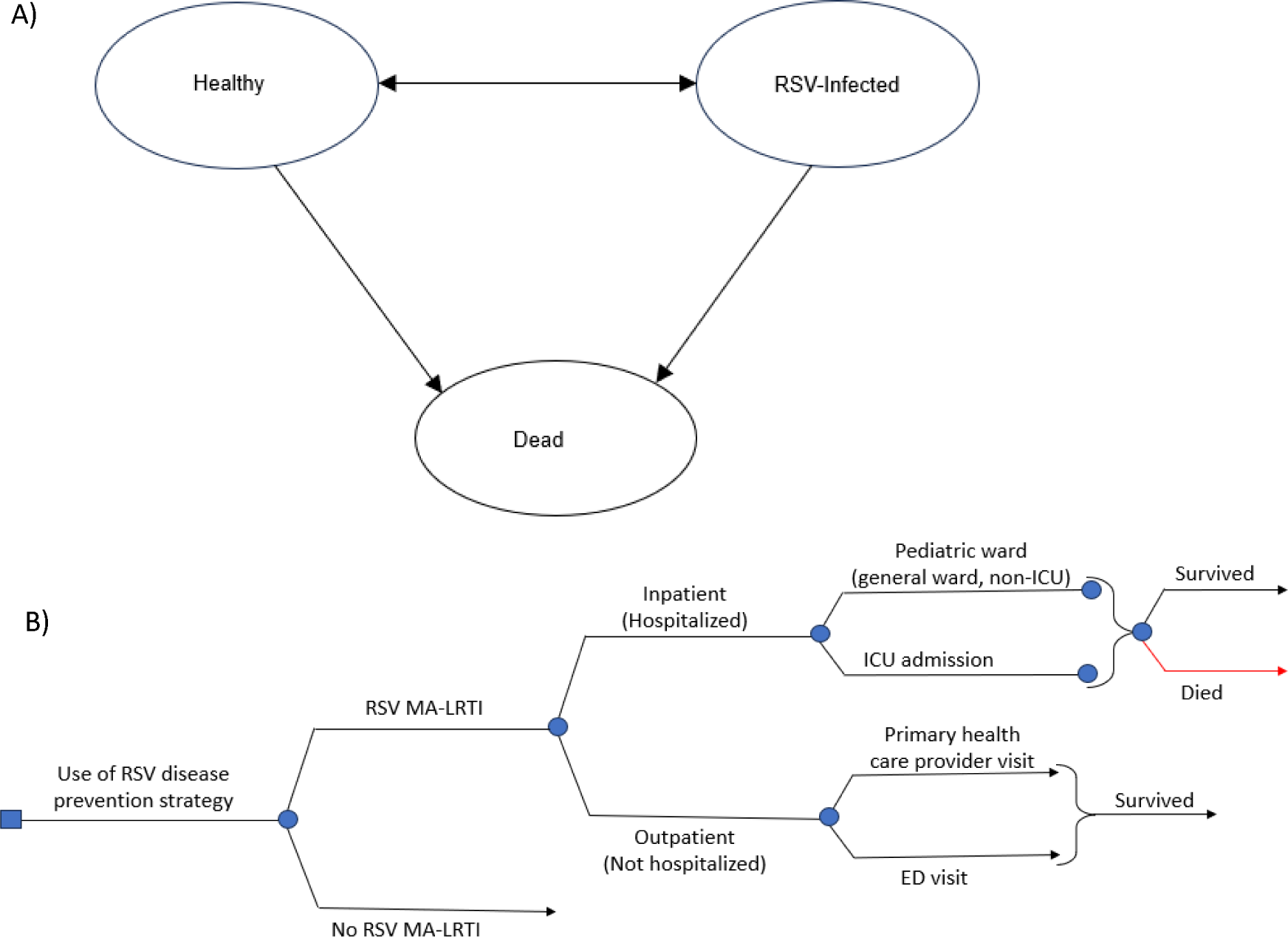
Overview of (A) health states and (B) structure of the model for RSV-related outcomes. Infants may transition to death from other causes not related to RSV disease (background mortality, arrows not shown). ED: emergency department; ICU: intensive care unit; MA-LRTI: medically-attended lower respiratory tract infection; RSV: respiratory syncytial virus

### Model parameters

Model parameters describing RSV disease epidemiology (Table 1), immunization product characteristics (Table 2), costs (Table 3), and QALY losses (Table 4) were obtained from available data and published studies, wherever possible, and by assumption otherwise. Canadian data were used preferentially.

**Table 1.** Epidemiological parameters.

| Parameter | Base | Range | Reference |
| --- | --- | --- | --- |
| Number of preterm births per 100 live births |  |  |  |
| <33 wGA | 1.2 |  | PHAC, 2020; Statistics Canada, 2023 (15, 16) |
| 33<sup>0</sup>-36<sup>6</sup> wGA | 6.8 |  |  |
| Monthly incidence of RSV infection per 1,000 infants |  |  |  |
| January | 28.55 |  | Rafferty *et. al.*, 2022 (14) |
| February | 36.65 |  |  |
| March | 30.97 |  |  |
| April | 13.06 |  |  |
| May | 6.34 |  |  |
| June | 2.61 |  |  |
| July | 0.87 |  |  |
| August | 0.70 |  |  |
| September | 0.00002 |  |  |
| October | 2.26 |  |  |
| November | 7.17 |  |  |
| December | 29.22 |  |  |
| RSV-attributable hospitalization, by gestational age (%) |  |  |  |
| <33 wGA | 5.1 | 4.0 – 6.3 | NACI, 2022 (17) |
| 33<sup>0</sup>–36<sup>6</sup> wGA | 3.3 | 2.7 – 4.1 |  |
| ≥37 wGA | 1.2 | 1.1 – 1.2 |  |
| ICU admission, from patients hospitalized with RSV (%) |  |  |  |
| <33 wGA | 51.7 | 41.3 – 62.1 | NACI, 2022 (17) |
| 33<sup>0</sup>–36<sup>6</sup> wGA | 31.5 | 13.1 – 53.6 |  |
| ≥37 wGA | 15.8 | 5.4 – 30 |  |
| Length of stay in pediatric ward (days) |  |  |  |
| <33 wGA | 6.1 | 0.48 – 12.71 | Banerji *et. al.*, 2016; Lanctôt *et. al.*, 2008; NACI, 2022 (17, 25, 27) |
| 33<sup>0</sup>–36<sup>6</sup> wGA | 3.9 | 0.64 – 6.08 |  |
| ≥37 wGA | 3.9 | 0.64 – 6.08 |  |
| Length of stay in ICU (days) |  |  |  |
| <33 wGA | 9.5 | 0.47 – 20.22 | Banerji *et. al.*, 2016; Lanctôt *et. al.*, 2008; NACI, 2022 (17, 25, 27) |
| 33<sup>0</sup>–36<sup>6</sup> wGA | 5.2 | 0.42 – 12.38 |  |
| ≥37 wGA | 5.2 | 0.42 – 12.38 |  |
| RSV mortality per hospitalized patient (30 days) (%) |  |  |  |
| 0-12 months (adjusted for background mortality from Statistics Canada (19) ) | 0.192 |  | Bourdeau *et. al.* 2023; Buchan *et. al.* 2023 (3, 18) |
| All-cause mortality per 1,000 population (per year) |  |  |  |
| 0 – 11 months | 4.33 |  | Statistics Canada, 2022 (19) |
| 12 – 24 months | 0.22 |  |  |

ICU: intensive care unit; PHAC: Public Health Agency of Canada; RSV: respiratory syncytial virus; wGA: weeks of gestational age

**Table 2.** Immunization products characteristics.

| Parameter | Base | Range | Reference |
| --- | --- | --- | --- |
| Immunization coverage (%) |  |  |  |
| RSVpreF | 64.8 | 52.4 – 80 | PHAC, 2023 (13) |
| Nirsevimab (among infants at moderate- and low-risk) | 71 |  | Kieffer *et. al.*, 2022 (12) |
| Nirsevimab (among infants at high-risk) | 80 |  |  |
| Palivizumab (among infants at high-risk) | 80 |  | Kieffer *et. al.*, 2022; Assumption (12) |
| Effectiveness (%) |  |  |  |
| RSVpreF |  |  |  |
| Effectiveness (0-5 months) against RSV-associated LRTI | 52.5 | 28.7 – 68.9 | Kampmann *et. al.*, 2023 (22) |
| Effectiveness (0-5 months) against RSV-associated hospitalization in pediatric ward and ICU | 56.4 | 5.2 – 81.5 |  |
| Effectiveness 6+ months | 0.0 | 0.0 | Assumption |
| Nirsevimab |  |  |  |
| Effectiveness (0-5 months) against RSV-associated LRTI | 80 | 70 – 87 | Griffin *et. al.*, 2020; Muller *et. al.*, 2023 (20, 21) |
| Effectiveness (0-5 months) against RSV-associated hospitalization | 81 | 64 – 90 |  |
| Effectiveness (0-5 months) against RSV-associated ICU admission | 90 | 54 – 98 |  |
| Effectiveness 6+ months | 0.0 | 0.0 | Assumption |
| Palivizumab |  |  |  |
| Effectiveness (0-1 month) against RSV-associated LRTI | 70 | 19 – 90 | Viguria *et. al.*, 2021 (23) |
| Effectiveness (0-1 month) against RSV-associated hospitalization | 82 | 29 – 96 |  |
| Effectiveness >1 month | 0.0 | 0.0 |  |
| Wastage rate (%) |  |  |  |
| RSVpreF | 5 | 0 – 25 | WHO, 2019 (28) |
| Nirsevimab | 5 | 0 – 25 |  |
| Palivizumab | 10 | 0 – 25 |  |

ICU: intensive care unit; PHAC: Public Health Agency of Canada; RSV: respiratory syncytial virus

**Table 3.** Cost parameters.

| Parameter | Base | Range | Reference |
| --- | --- | --- | --- |
| Cost of administration per dose (\$) | 14.7 | 0 – 30 | Papenburg *et. al.*,2020; Abu-Raya *et. Al.*,2020 (32, 60) |
| Cost per product dose (\$) | | | |
| RSVpreF | 230 | 180 – 400 | Robertson, 2023 (61) |
| Nirsevimab | 952 | 180 – 1000 | Public listed price, Sanofi |
| Palivizumab | 1,227 | 700 – 2000 | INESSS, 2017; Smart *et. al.*, 2010; Assumption (range) (29, 30) |
| Cost per patient with RSV managed in inpatient setting per day (\$) | | | |
| Per diem in pediatric general ward | 1,491 | 1118 – 1864 * | Lancôt *et. al.*, 2008 (25) |
| Per diem in ICU | 3,638 | 2329 – 4548* | Lancôt *et. Al.*, 2008; CIHI, 2016 (25, 26) |
| Attributable 30-day cost per patient with RSV requiring outpatient healthcare provider visit (\$) | | | |
| <3 months | 245 | 184 – 306 * | Rafferty *et. al.*, 2022 (14) |
| 3 - <6 months | 136 | 102 – 170* |  |
| 6m - <1 year | 133 | 100 – 166 * |  |
| 1- <2 years | 120 | 90 – 150* |  |
| Attributable 30-day cost per patient with RSV requiring ED visit (\$) | | | |
| <3 months | 225 | 169 – 281 * | Rafferty *et. al.*, 2022 (14) |
| 3 - <6 months | 131 | 98 – 164 * |  |
| 6m - <1 year | 120 | 90 – 150* |  |
| 1- <2 years | 112 | 83 – 140* |  |
| Out-of-pocket costs (\$) | | | |
| Cost of transportation to vaccination | 3.66 | 2.75 – 4.58* | Mitchell *et. al.*, 2017 (36) |
| Cost of transportation to outpatient care | 3.66 | 2.75 – 4.58* |  |
| Cost of transportation to inpatient care | 182 | 14 – 1,925 |  |
| Cost of childcare and home health after inpatient discharge | 329 | 120 – 963 |  |
| Other out-of-pocket costs for patients with RSV managed in inpatient setting (such as over-the-counter medications and other non-transportation or home expenses) | 375 | 36 – 5,239 |  |
| Caregiver workdays lost |  |  |  |
| Hospitalization | 7.3 | 1.125 – 68.1 | Mitchell *et. al.*, 2017 (36) |
| Outpatient healthcare provider visit | 2.5 | 0.5 – 5 | Fragaszy *et. al.*, 2018 (37) |
| ED visit | 2.5 | 0.5 – 5 |  |
| Visit healthcare provider for vaccination | 0.5 |  | Assumption |
| Labour force participation (%) |  |  |  |
| Age 16+ | Age-specific values |  | Statistics Canada, 2022 (34) |
| Caregiver | 88.6% |  | Statistics Canada, 2022 (34) |
| Average employment income (\$) | | | |
| Age 16+ | Age-specific values |  | Statistics Canada, 2022 (35) |
| Caregiver | 62,400 |  | Statistics Canada, 2022 (35) |

\*Range defined as $\pm 25\%$ of the base value
ED: emergency department; ICU: intensive care unit; RSV: respiratory syncytial virus

### RSV disease epidemiology

Estimates of monthly incidence of RSV infection were based on a study for the province of Alberta, which provided projection of RSV cases from 2010 to 2019 (14). RSV seasonality was assumed to have returned to pre-pandemic seasonal patterns, with peak incidence occurring in December to March [*PHAC surveillance; RVDSS RSV surveillance data, 2023 (Personal communication)*]. The proportion of preterm births was obtained from Statistics Canada (15, 16). In Canada, extremely/very preterm infants accounted for 1.2% of all live births and 6.8% of live births were late preterm infants (15, 16). The rates of RSV-associated hospitalizations by gestational age at birth were derived from a systematic review (17). Case-fatality and age-specific background mortality rates were obtained from the literature (3, 18, 19). The case-fatality rate was adjusted to account for background mortality.

### Effectiveness of monoclonal antibodies and vaccine

Effectiveness and duration of protection data were obtained from targeted literature searches. A meta-analysis of two randomized controlled trials (RCTs) estimated the pooled effectiveness of a single weight-banded dose of nirsevimab to be 80% against RSV MA-LRTI, 81% in reducing RSV-associated hospitalization and 90% against very severe RSV disease (defined as RSV-associated ICU admission in this analysis) for the first 150 days post administration (20, 21). The effectiveness of nirsevimab was assumed to drop to 0% after 150 days. RSVpreF vaccine effectiveness against RSV MA-LRTI and hospitalization during the first 150 days of life was estimated at 52.5% and 56.4%, respectively (22). It was assumed that the RSVpreF vaccine had the same effectiveness among full-term (low-risk) and late preterm (moderate-risk) infants. The effectiveness of RSVpreF assumed to drop to 0% after 150 days. RSVpreF was assumed to have no effect on infants born before 32 wGA. The analysis did not include RSVpreF effectiveness for preventing RSV disease in pregnant women and pregnant people as no data exist. A single dose of palivizumab was assumed to be 70% and 80% effective in preventing RSV MA-LRTI and hospitalization in infants at high-risk, respectively (23). It was also assumed that a single dose of palivizumab would provide one-month of protection, with infants receiving five doses during their first RSV season (24).

### Costs

Costs included immunization costs and RSV-associated medical expenses for outpatient healthcare provider visits, ED visits, hospitalization in a pediatric general ward, and ICU. Pediatric general ward hospitalization and ICU costs were calculated using the per diem costs (25, 26) with a corresponding length of hospital stay derived from literature (17, 25, 27). Outpatient healthcare provider and ED visit costs were obtained from a population-based matched retrospective case–control study using administrative data from Alberta (14). The costs of immunization included administration cost, product price, and wastage costs (28). Canadian list prices were used in the base case for nirsevimab (both 50 and 100 mg vial) and RSVpreF: $952 and $230 per dose, respectively. Wastage costs were calculated as per the World Health Organization (WHO) recommendations (28). The weighted average price of palivizumab was calculated to be $1,227 per dose, with 36.96% of infants receiving the 50 mg vial at a cost of $752 per dose and 63.04% receiving the 100 mg vial for $1,505 per dose (25, 29–31). The cost of administration for vaccine or monoclonal antibodies was set to $14.70 per dose, with each administration considered a new visit (32). Cost of adverse events following immunization were not included in the model. All costs are in 2023 Canadian dollars and, where necessary, adjusted using the Canadian Consumer Price Index (33).

For the societal perspective, costs included productivity loss due to death from RSV disease, caregiver costs, and out-of-pocket costs (i.e., transportation, over-the-counter medications, home expenses). Productivity loss was estimated using the human capital method. Age-specific labour force participation rates (34) and average employment income (35) were obtained from Statistics Canada. Caregiver days lost due to hospitalization were assumed to be 7.3 days (36). Caregiver days lost for outpatient and ED visits was assumed to be 2.5 days based on estimates for adults with acute respiratory tract infection (37). Caregiver wages were calculated based on the average employment income (35) and labour force participation of the population aged 25 to 54 years old (34).

### QALY losses

RSV-associated QALY loss was assessed for both infants and their caregivers. The disutility weights for hospitalized infants and their caregivers (one per child) were derived from a systematic review, which compared the decrease in utility of RSV-hospitalized infants to those hospitalized for other reasons (38). There was a 45% increase in utility loss for infants admitted to the ICU and their caregivers, compared to those hospitalized in a pediatric general ward, based on observed utility difference for infants with these different RSV outcomes (39). Utility decrements for outpatient healthcare provider or ED visits for RSV were derived from a previous cost-effectiveness study of RSV prophylactic products that estimated QALY loss based on pertussis (40). Caregiver QALY losses were about 50% of children’s losses (38).

### Cost-effectiveness analysis

Deterministic model estimates were calculated for a cohort of 1,000 infants. A sequential analysis was conducted to compare multiple RSV disease prevention strategies. Sequential ICERs were calculated by ordering the strategies from lowest to highest cost and comparing incremental costs and QALYs gained for a given strategy to the next less costly strategy. A strategy was eliminated if there were other strategies that were projected to result in more QALYs gained at lower costs (i.e., the strategy is dominated) or there was a combination of other strategies that would result in more QALYs gained for lower costs, such that the excluded strategy would never be the optimal intervention, regardless of the cost-effectiveness threshold used (i.e., the strategy was subject to extended dominance). Unless specified otherwise, results are presented for the health system perspective, with results for the societal perspective provided in the supplementary material.

Sensitivity of the results to individual model parameters was examined in a one-way sensitivity analysis, where each parameter was varied one at a time over the ranges listed in Table 1 to Table 4. Given the uncertainty in per dose prices of RSVpreF and nirsevimab, a two-way sensitivity analysis was conducted to assess the combined impact of varying the prices of both RSVpreF and nirsevimab. Results are presented at cost-effectiveness thresholds of $50,000 and $100,000 per QALY.

**Table 4.**
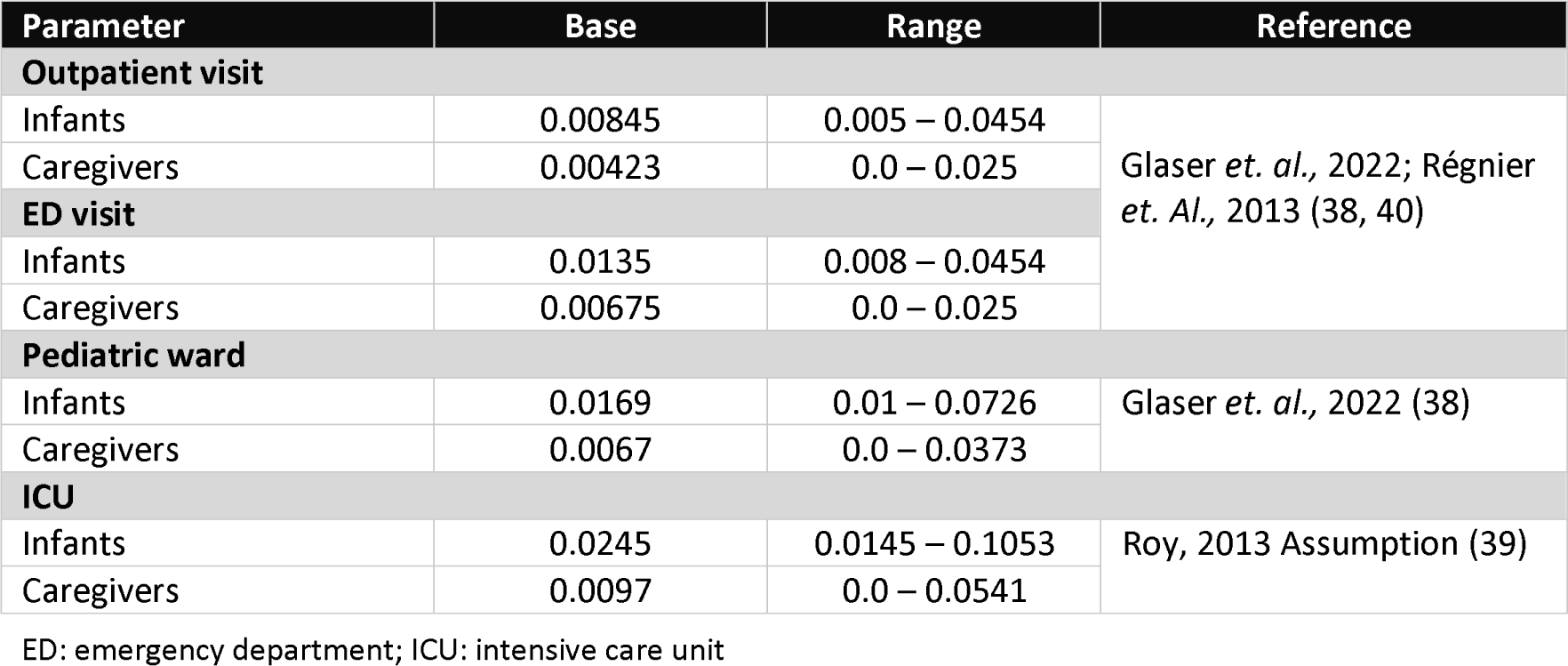
RSV-associated QALY losses.

Scenario analyses were conducted to explore the potential impact of longer duration of protection of nirsevimab and/or RSVpreF. For these analyses, the effectiveness of both nirsevimab and RSVpreF was assumed to remain constant for the first 5 months as estimated in published trials (22, 41), followed by a linear decline to no protection after 10 and 8 months, respectively.

Considering the significant productivity loss incurred by the caregivers during the administration of the vaccine or monoclonal antibodies ($141 per visit), a scenario analysis was conducted from the societal perspective assuming no caregiver costs when they visit the healthcare facility for immunization.

A scenario analysis was also performed to evaluate the impact of the interventions in settings with higher hospitalization rates and higher healthcare costs, which could better reflect realities for some communities, such as in Northern Canada. Infants in this setting were assumed to have a five-fold higher rates of RSV hospitalization than the rest of Canada (42, 43). The average cost per patient requiring outpatient care, pediatric general ward hospitalization, and ICU admission were estimated to be $1,747,

$18,869, and $73,532, respectively (44). Cost of transportation (including medical evacuation cost of $16,576) was $18,010 (45), and administration cost for each dose was $50 (44, 46). These parameter values were applied to the base case model population (reflective of the entire Canadian population) to provide a generalized comparison of differences in cost-effectiveness relative to the base case results that could be applicable to jurisdictions across Canada experiencing higher RSV-associated hospitalizations and costs.

## RESULTS

### Base case analysis

The number of RSV-related health outcomes averted by RSVpreF and nirsevimab programs compared to palivizumab for infants at high-risk program is depicted in Figure 2. All of the all-infants nirsevimab programs that were modeled prevented more cases of RSV-related outcomes than year-round RSVpreF over the model time period. Of all interventions considered, seasonal nirsevimab programs for all infants with catch-up averted the most RSV-related outcomes, while year-round RSVpreF alone prevented the fewest RSV outcomes. For instance, a seasonal nirsevimab program for all infants with catch-up prevented 51 additional outpatient healthcare provider visits for RSV per 1,000 population over the 1-year study period, compared to 20 cases avoided by a year-round RSVpreF program.

**Figure 2.**
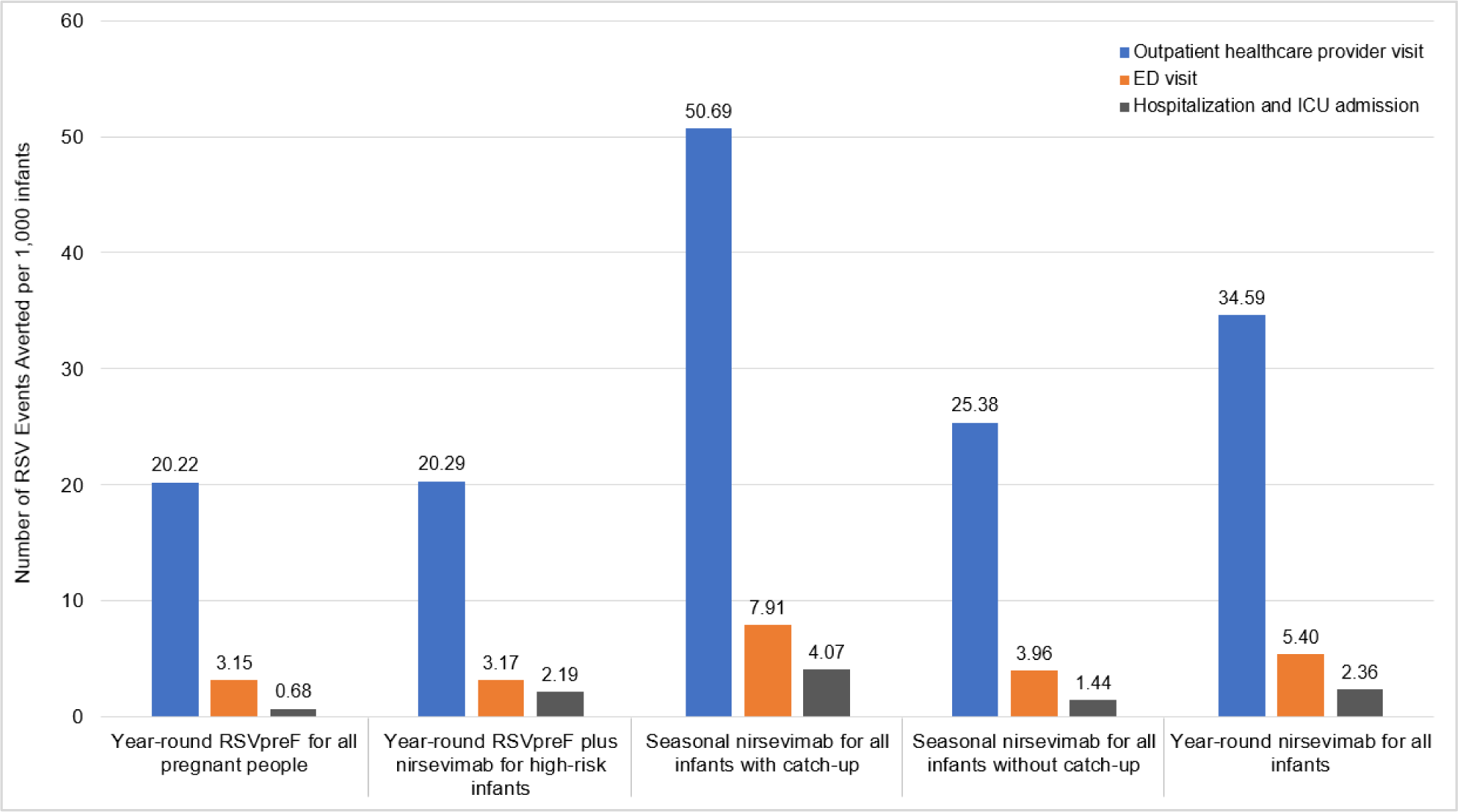
RSV-related health outcomes averted in infants with the use of year-round RSVpreF and all-infants nirsevimab programs compared to palivizumab standard of care. Results are based on deterministic analysis for a cohort of 1,000 infants. **ED: emergency department;ICU: intensive care unit; RSV: respiratory syncytial virus**

Figure 3 and Supplementary Table 1 shows results of the analysis in which all interventions are compared sequentially against each other and the standard of care, from the health system perspective (societal ICERs are provided in supplementary Figure 2). All strategies were dominated, except seasonal nirsevimab programs with catch-up (either for all infants or restricted to infants at moderate- or high-risk), and year-round RSVpreF for all pregnant women and people plus nirsevimab for infants at high-risk. Of all the interventions, seasonal nirsevimab for infants at moderate- and high-risk with catch-up was the most cost-effective strategy with an ICER of $27,891 per QALY when compared to palivizumab standard of care. The ICER for year-round RSVpreF for all pregnant women and pregnant people plus nirsevimab for infants at high-risk compared to seasonal nirsevimab for infants at moderate- and high-risk with catch-up was $204,621 per QALY. A seasonal nirsevimab program for all infants with catch-up resulted in a higher ICER ($512,265 per QALY) when compared to year-round RSVpreF plus nirsevimab for infants at high-risk.

**Figure 3.**
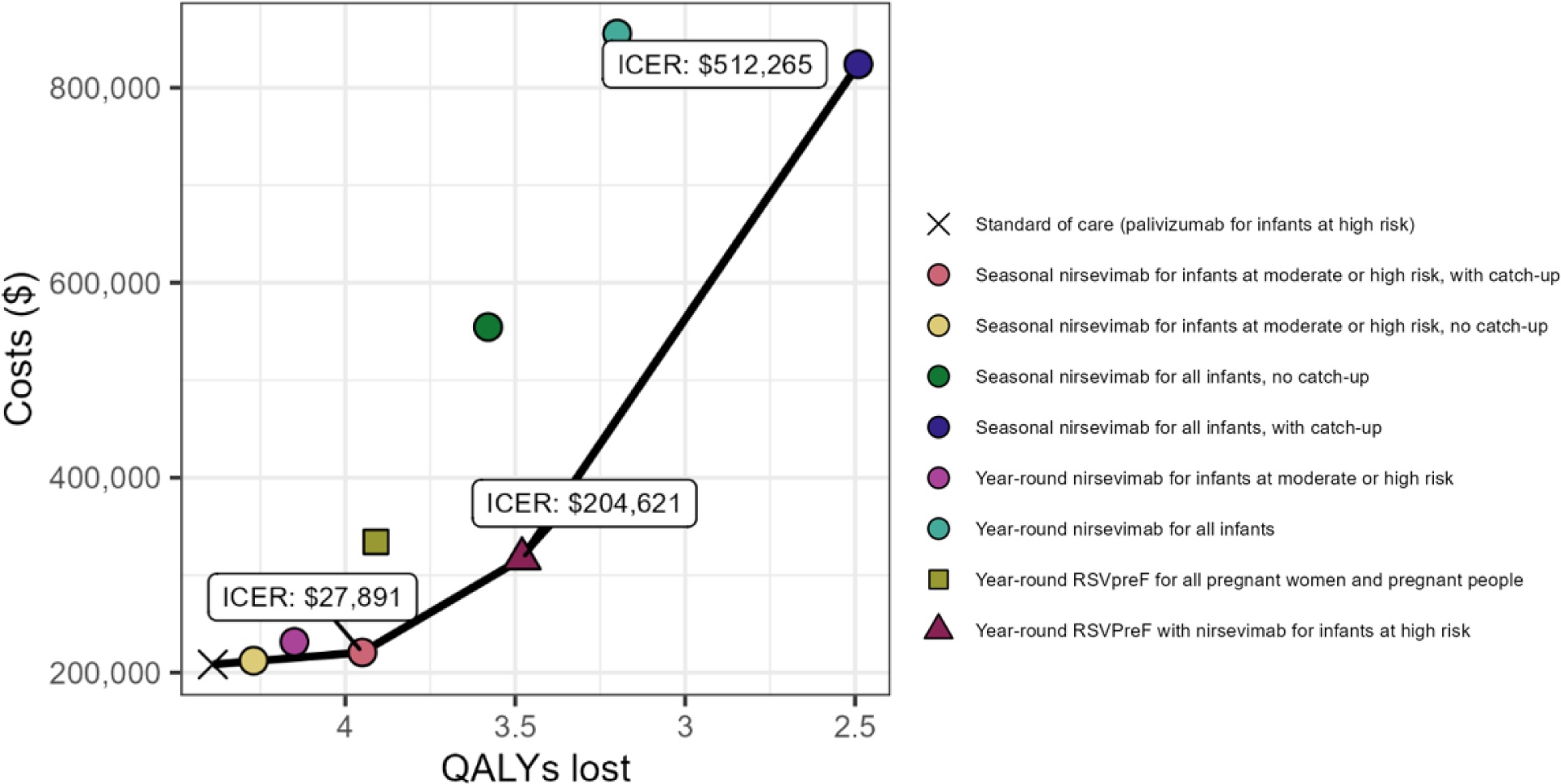
Base case results of sequential analysis comparing all RSV disease prevention strategies against each other and standard of care from the health system perspective. Note that the seasonal nirsevimab program for infants at moderate- and high-risk without catch-up was dominated, but the dot representing this program appears to be on the efficiency frontier due to the scale of the graph.

### Sensitivity and scenario analyses

In one-way sensitivity analyses, ICERs for seasonal nirsevimab for infants at moderate- and high-risk with catch-up compared to palivizumab were most sensitive to changes in nirsevimab price, medical costs for infants at high-risk with RSV managed in the ICU, nirsevimab effectiveness against ICU admission, palivizumab effectiveness against hospitalization, and RSV monthly infection rates (Figure 4). Similarly, per dose price of nirsevimab and RSVpreF, RSVpreF effectiveness against MA-LRTI, RSV-associated QALY loss, and RSV monthly infection rates were found to be influential parameters for RSVpreF plus nirsevimab for infants at high-risk program compared to a seasonal nirsevimab program for infants at moderate- and high-risk. In this analysis, the ICER only fell below $100,000 per QALY if RSV-associated QALY loss was higher than assumed in the base case. Compared to RSVpreF plus nirsevimab for infants at high-risk, a seasonal nirsevimab program for all infants with catch-up was never cost-effective within the range of values evaluated (not shown).

**Figure 4.**
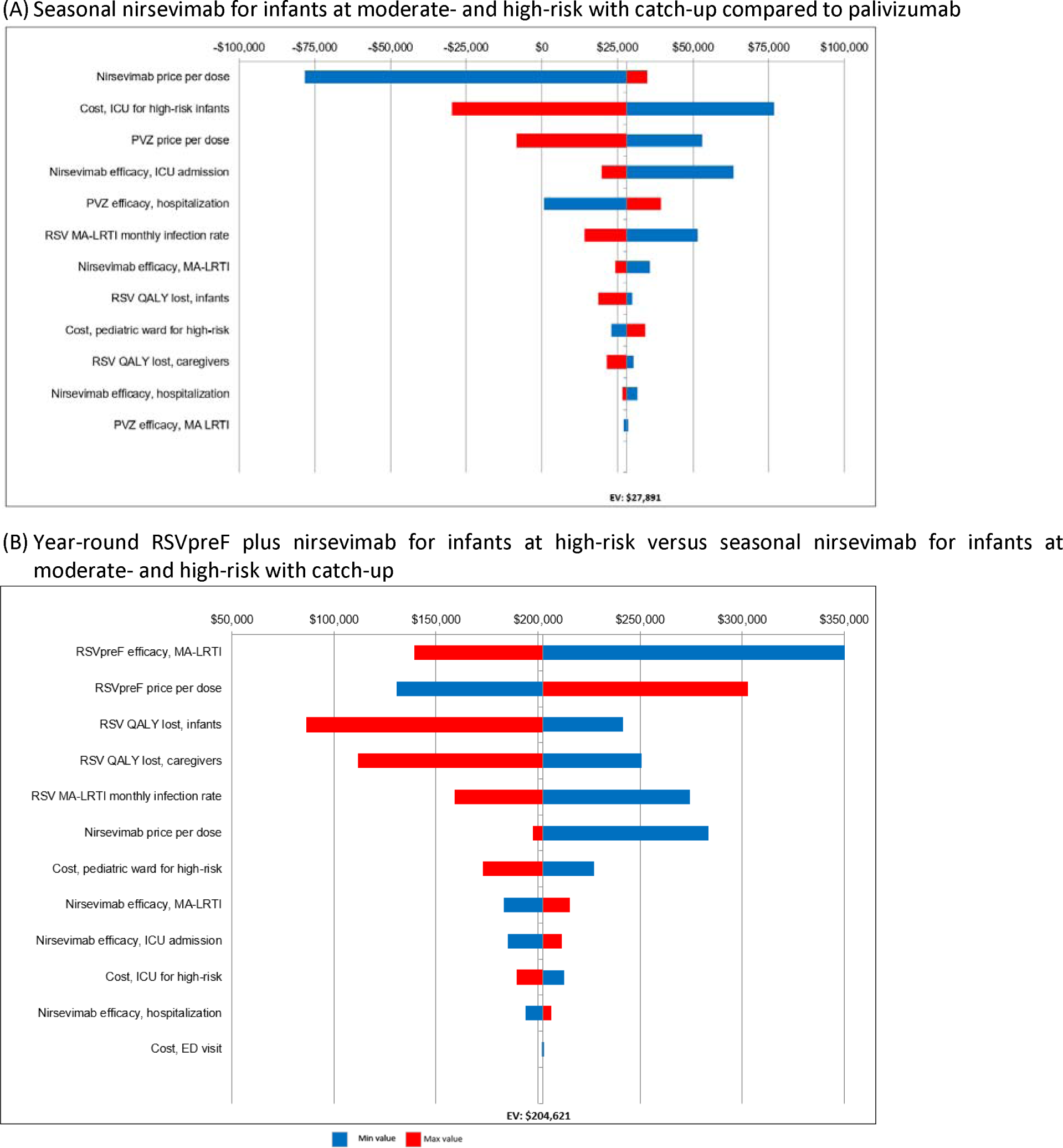
One-way sensitivity analyses of key parameters comparing: (A) seasonal nirsevimab for infants at moderate- and high-risk with catch-up versus palivizumab program; (B) year-round RSVpreF plus nirsevimab for infants at high-risk versus seasonal nirsevimab for infants at moderate- and high-risk with catch-up (base-case ICER of $ 204,621 per QALY). Note that a negative ICERs signifies that the interventions are dominant, meaning they are less costly and more effective than the comparator. **ED: emergency department; ICU: intensive care unit; MA-LRTI: medically-attended lower respiratory tract infection; QALY: quality-adjusted life years; PVA: palivizumab; RSV: respiratory syncytial virus**

The combined impact of varying the prices of both RSVpreF and nirsevimab on base case results was examined in a two-way sensitivity analysis under different cost-effectiveness thresholds ($50,000 or $100,000 per QALY), with nirsevimab priced at $0-1,000 and RSVpreF priced $0-400 (Figure 5). When all strategies were compared sequentially against each other and the standard of care, seasonal nirsevimab for infants at moderate- and high-risk with catch-up was found to be an optimal strategy, with the following exceptions: seasonal nirsevimab for all infants with catch-up was optimal if the price of nirsevimab was <$110-290; and year-round RSVpreF plus nirsevimab for infants at high-risk would be cost-effective when the price of nirsevimab was >$110-290 and the price of RSVpreF was <$60-155. Seasonal nirsevimab without catch-up, year-round nirsevimab and year-round RSVpreF programs were never cost-effective options at commonly used thresholds.

**Figure 5.**
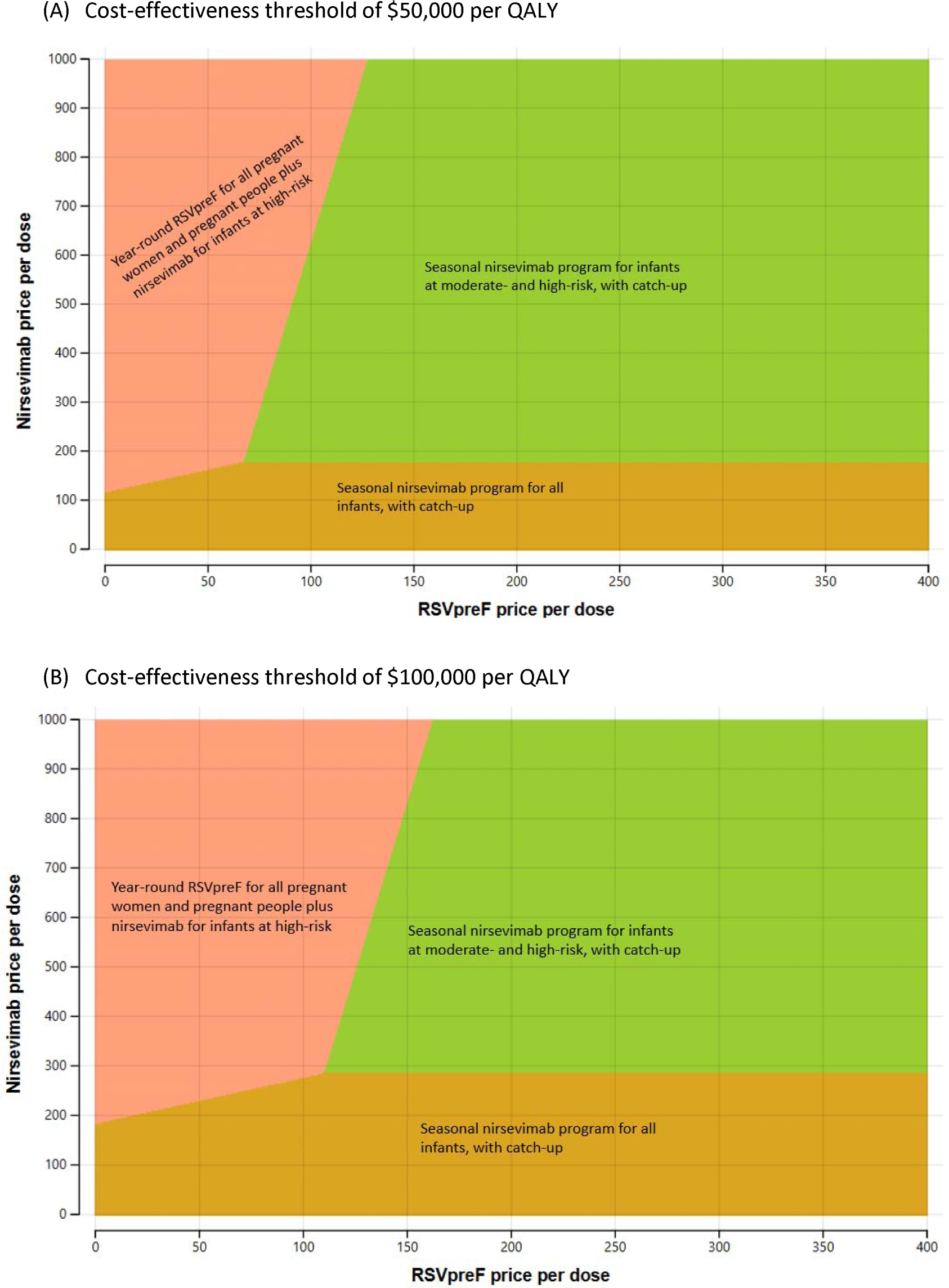
Two-way sensitivity analyses by varying prices of nirsevimab and RSVpreF when comparing all strategies including standard of care: (A) at a cost-effectiveness threshold of $50,000 per QALY; (B) at a cost-effectiveness threshold of $100,000 per QALY. The optimal strategy is the colored tile corresponding to the programs.

While the extended duration of nirsevimab protection resulted in lower ICERs for nirsevimab programs, nirsevimab for all infants strategies were still unlikely to be considered cost-effective (Figure 6). A year-round RSVpreF program was dominated even when duration of protection of RSVpreF was assumed to extend beyond the five months used in the base case analysis. When longer durations of protection for nirsevimab and/or RSVpreF were assumed, ICERs for RSVpreF plus nirsevimab for infants at high-risk remained >$132,000 QALY.

**Figure 6.**
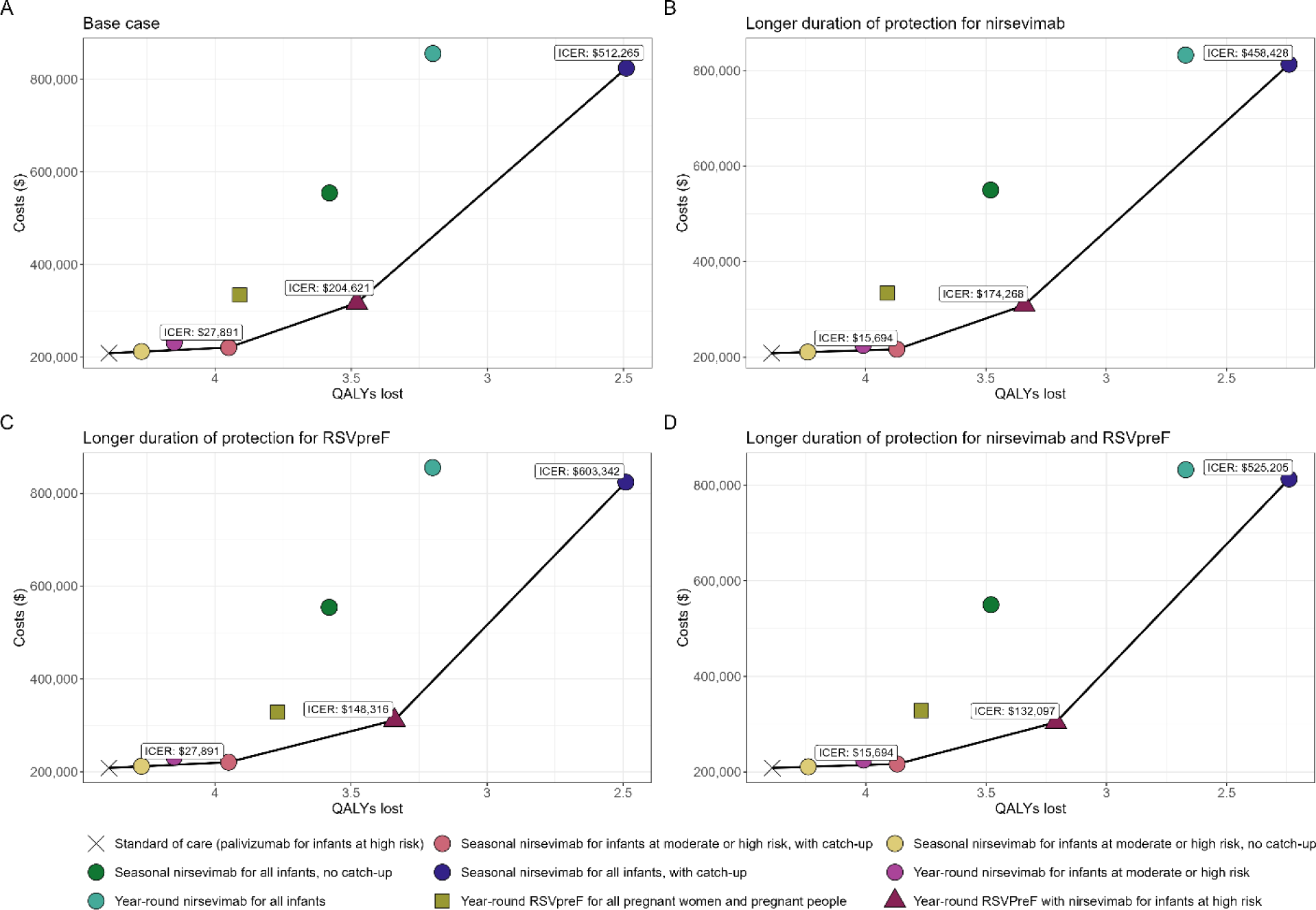
Impact of longer duration of protection for nirsevimab and/or RSVpreF, when all strategies compared sequentially against each other and standard of care. ICERs are presented for (A) Base case results; (B) Longer duration of protection for nirsevimab; (C) Longer duration of protection for RSVpreF; (D) Longer duration of protection for both nirsevimab and RSVpreF.

A scenario analysis assuming no caregiver costs during the administration of the vaccine or monoclonal antibodies from the societal perspective resulted in lower ICERs for nirsevimab and RSVpreF programs, but the overall conclusions remained unchanged (Supplementary Figure 3).

Results for settings with higher RSV burden and higher health care costs are summarized in Figure 7. When comparing all strategies sequentially, a seasonal nirsevimab program for all infants with catch-up was dominant (less costly and more effective), except for the year-round RSVpreF for all pregnant women and pregnant people plus nirsevimab for infants at high-risk. An all-infants seasonal nirsevimab program with catch-up had an ICER of $5,700 per QALY when compared to a combined program of year-round RSVpreF offered to all pregnant women and pregnant people followed by a year-round nirsevimab for infants at high-risk. For the societal perspective, a seasonal nirsevimab for all infants with catch-up dominated all other strategies considered.

**Figure 7.**
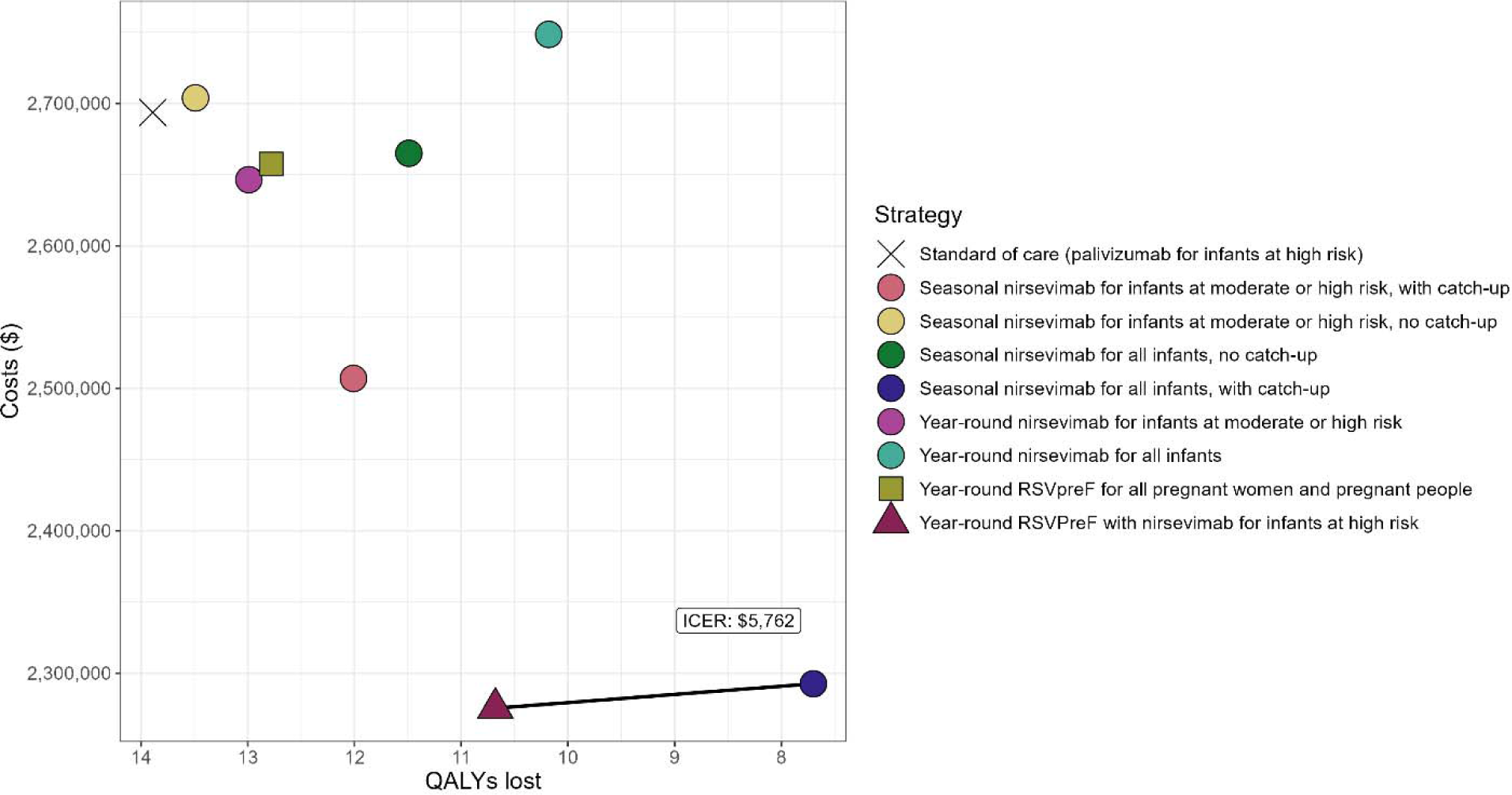
Cost-effectiveness frontiers for a scenario with higher RSV hospitalization rate and health care costs, when comparing all strategies including standard of care from the health system perspective.

## DISCUSSION

This study evaluated the cost-effectiveness of various RSVpreF and nirsevimab immunization strategies for protecting infants against RSV disease in Canada. We found that all modelled RSVpreF and nirsevimab programs prevented additional cases of RSV disease than the current palivizumab program, with nirsevimab programs for all infants averting more health events than RSVpreF programs. A year-round RSVpreF program had lower intervention costs than all-infants nirsevimab programs due to the lower per dose price of RSVpreF and lower assumed vaccination coverage, but its lower effectiveness and coverage led to higher RSV-related costs.

Our results are comparable to studies from high-income countries including Canada (47, 48), United States (49–52), England and Wales (53), Norway (54), six European countries (55) and the findings from two systematic reviews conducted by Canadian Agency for Drugs and Technologies in Health (CADTH) (56, 57). The ICER values varied considerably mainly due to differences in key parameter inputs (such as vaccine effectiveness, product prices, and RSV burden), model structure, and type/number of strategies compared including the comparator, underscoring the sensitivity of the results to model assumptions and inputs. However, these studies demonstrated that all-infants nirsevimab programs alone or in combination with pregnancy vaccines (which may not be specific to RSVpreF as several studies compared theoretical maternal vaccines) generated ICERs that generally exceeded commonly accepted cost-effectiveness thresholds. Our study also showed that a seasonal nirsevimab program with catch-up was cost-effective only when given to preterm infants born before 37 wGA or infants living in communities with higher RSV burden and healthcare costs (reflective of complex transport). Similar to our study, several studies demonstrated that a seasonal nirsevimab program with catch-up dominated year-round nirsevimab or a pregnancy vaccine (48, 49, 53, 55). A seasonal nirsevimab program with catch-up was more effective in reducing RSV burden than a year-round program as it ensures protection when infants require it most (i.e., during the RSV season) rather than providing protection outside the season. However, the effectiveness and cost-effectiveness of a seasonal program assumes that we know the start and end of the RSV season, which may be challenging, as demonstrated by disruptions in RSV seasonality associated with the COVID-19 pandemic (58, 59). The sensitivity of seasonal programs to timing of the RSV season underscores the importance of robust surveillance systems to determine optimal timing for implementing seasonal interventions.

Our price threshold analysis indicated that an all-infants seasonal nirsevimab program with catch-up could be cost-effective if nirsevimab price was reduced by at least 80-88% (at a cost-effectiveness threshold of $50,000 per QALY). A year-round RSVpreF program alone was never a cost-effective option across analyses, but use of year-round RSVpreF in combination with nirsevimab for infants at high-risk could be an optimal strategy (at a threshold of $50,000 per QALY) if the price of nirsevimab was reduced by less than 12-20% of the list price and the price of RSVpreF was reduced by more than 45-74%. Similarly, previous studies reported that nirsevimab and RSVpreF prices need to be substantially lowered in order for them to be considered cost-effective (48, 50, 53, 55). For instance, a Canadian study found that year-year-round RSVpreF program may be cost-effective if the price of RSVpreF is below $160, while that of nirsevimab program for all infants may be cost-effective if price of nirsevimab is below $215 per dose (48).

We did not evaluate the impact of seasonal RSVpreF programs due to anticipated implementation and feasibility challenges, but studies from other jurisdictions reported a lower ICER for seasonal RSVpreF program than a year-round (51, 53). A static model by the United States Centers for Disease Control and Prevention estimated an ICER for seasonal RSVpreF (administered between September and January) of $227,000 (US$167,280)) per QALY when compared to no vaccination versus $543,200 (US$400,304) per QALY for a year-round program in the base case, but ICERs remained above commonly used thresholds (51). A price threshold analysis using £20,000 per QALY threshold in the England and Wales study showed that a seasonal RSVpreF (administered between July and December) could be the optimal strategy if cost per dose and administration is less than $137 (£80) versus $60 (£35) per dose for a year-round program (53).

Our cost-effectiveness analysis of RSV prevention strategies for infants living in areas with higher hospitalization rates and healthcare expenses provides contextual insight into areas with complex medical transport, acknowledging that these areas are diverse and many have unique challenges that may not be captured by modelling. Our findings align with the study in Nunavik in northern Quebec where infants with severe RSV disease require transport to hospitals south of the region for medical care. The study found that administering a long-acting monoclonal antibody (nirsevimab) to all infants (i.e., healthy and high-risk infants) was cost-effective compared to no intervention, with ICERs ranging from $5,255 to $39,414 per QALY depending on the severity of the RSV season. The study also found that nirsevimab programs for infants at high-risk were dominant compared to no intervention, regardless of the severity of the RSV season. Comparable to our findings, RSVpreF alone was not cost-effective strategy (ICER of $227,286 per QALY) during mild RSV seasons (i.e., 30-50% of households had individuals infected with RSV) (47). Conversely, RSVpreF alone was dominant in moderate to severe RSV seasons (i.e., ≥50% of households had individuals infected with RSV) (47).

Although we developed a comprehensive model of medically-attended RSV infection and extensively explored parameter uncertainty, our study has limitations. The per dose price of RSVpreF and/or nirsevimab was a key driver of the model results and using public list prices (which are often higher than negotiated prices) may have led to overestimation of ICERs. However, our price threshold analysis identified the prices at which different strategies could be cost-effective at commonly used thresholds and this may offer valuable insights for decision-making. We used a static model that did not account for intervention effects on disease transmission; currently there is no evidence regarding indirect effects among unimmunized populations. While we would not expect the consideration of herd immunity to appreciably change our estimates given the relatively short duration of protection conferred by these interventions, future economic assessments should incorporate transmission dynamics if relevant data become available. Further, we did not model protection against upper respiratory tract infections, asymptomatic LRTI, and the potential long-term consequences of RSV disease associated with recurrent wheezing and asthma because the impact of the interventions on these outcomes remains uncertain.

Our model did not include costs and QALY losses associated with potential adverse events following immunization; however, given the expected rarity of serious adverse events, their inclusion would likely have minimal impact on the overall estimates. Moreover, we conducted deterministic analysis; however, performing probabilistic sensitivity analysis would have provided a more comprehensive understanding of the uncertainty in our results.

Some model assumptions may be refined over time. The model assumed that antibody-induced protection from the new passive immunizing strategies drops to 0% after 150 days. It may be more likely that protection would wane according to a gradient as antibody levels gradually decline, but the decay kinetics are not yet known so it was not possible to model this reduction more precisely. Although it was not considered feasible in the Canadian program environment, it is likely that a scenario including seasonal administration of RSVpreF, reflecting recent program design in the United States, would offer improved cost-effectiveness over a year-round RSVpreF program. This scenario was not modelled, but could be an area of future investigation.

### Summary

Use of RSVpreF vaccine or nirsevimab in an infant’s first season of RSV could significantly reduce the burden of RSV disease in this population. Nirsevimab programs were cost-effective when limited to infants born before 37 wGA or those residing in areas with higher RSV burden and healthcare costs, reflective of remote communities where transport for treatment of severe RSV disease would be complex. For all-infant programs to be cost-effective, a substantial reduction in product prices is required.

## Authors’ statement

GBG – Conceptualization, Modelling, Analysis, Manuscript drafting

MWY – Conceptualization, Analysis, Manuscript review and editing

RX – Conceptualization, Manuscript review and editing

AC – Conceptualization, Manuscript review and editing

AES – Conceptualization, Manuscript review and editing

AK – Conceptualization, Manuscript review and editing

WS – Conceptualization, Manuscript review and editing

ER – Conceptualization, Manuscript review and editing

NB – Conceptualization, Manuscript review and editing

MT – Conceptualization, Manuscript review and editing

ART – Conceptualization, Analysis, Manuscript review and editing

## Data Availability

All data produced in the present work are contained in the manuscript

## Acknowledgements

The authors would like to thank the NACI Secretariat team (Pamela Doyon-Plourde and Phaedra Davis), and the NACI RSV Working Group.

