## Supplementary Material for "Cost-effectiveness of RSVpreF vaccine and nirsevimab for the prevention of respiratory syncytial virus disease in Canadian infants"

**Supplementary Table 1**. Base case: mean costs, quality-adjusted life years lost, and sequential incremental cost-effectiveness ratios for nirsevimab and RSVpreF programs compared to a palivizumab program from the health system perspective*

| **Strategy** | **Costs**  **($)** | **Effect**  **(QALYs lost)** | **Sequential ICERs**  **($ per QALY)** |
| --- | --- | --- | --- |
| Standard of care (palivizumab for infants at high risk) | 208,527 | 4.39 | -- |
| Seasonal nirsevimab for infants at moderate or high risk, no catch-up | 212,251 | 4.27 | Dominated |
| Seasonal nirsevimab for infants at moderate or high risk, with catch-up | 220,799 | 3.95 | 27,891 |
| Year-round nirsevimab for infants at moderate or high risk | 231,567 | 4.15 | Dominated |
| Year-round RSVpreF plus nirsevimab for infants at high-risk | 316,971 | 3.48 | 204,621 |
| Year-round RSVpreF for all pregnant women and pregnant people | 334,187 | 3.91 | Dominated |
| Seasonal nirsevimab for all infants, no catch-up | 554,487 | 3.58 | Dominated |
| Seasonal nirsevimab for all infants, with catch-up | 824,113 | 2.49 | 512,265 |
| Year-round nirsevimab for all infants | 855,494 | 3.20 | Dominated |

^*^Results calculated for a cohort of 1,000 infants.

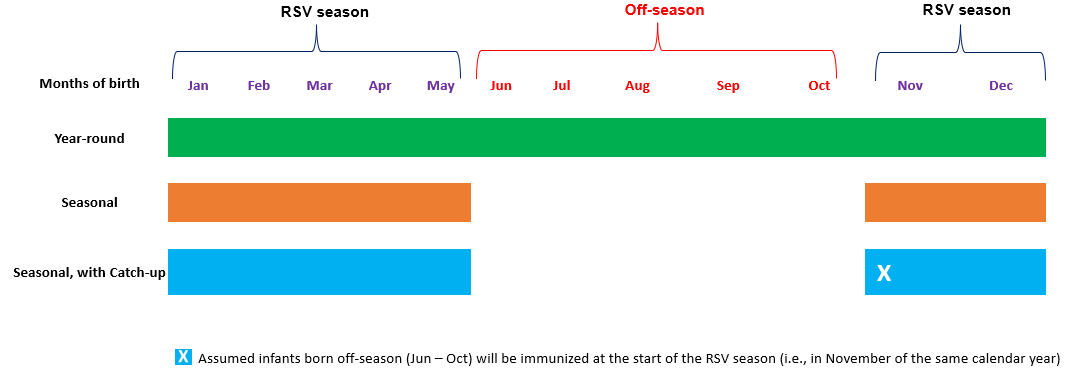

**Supplementary Figure 1:** Timing of seasonal and year-round administration of immunizing agents for the first season of RSV

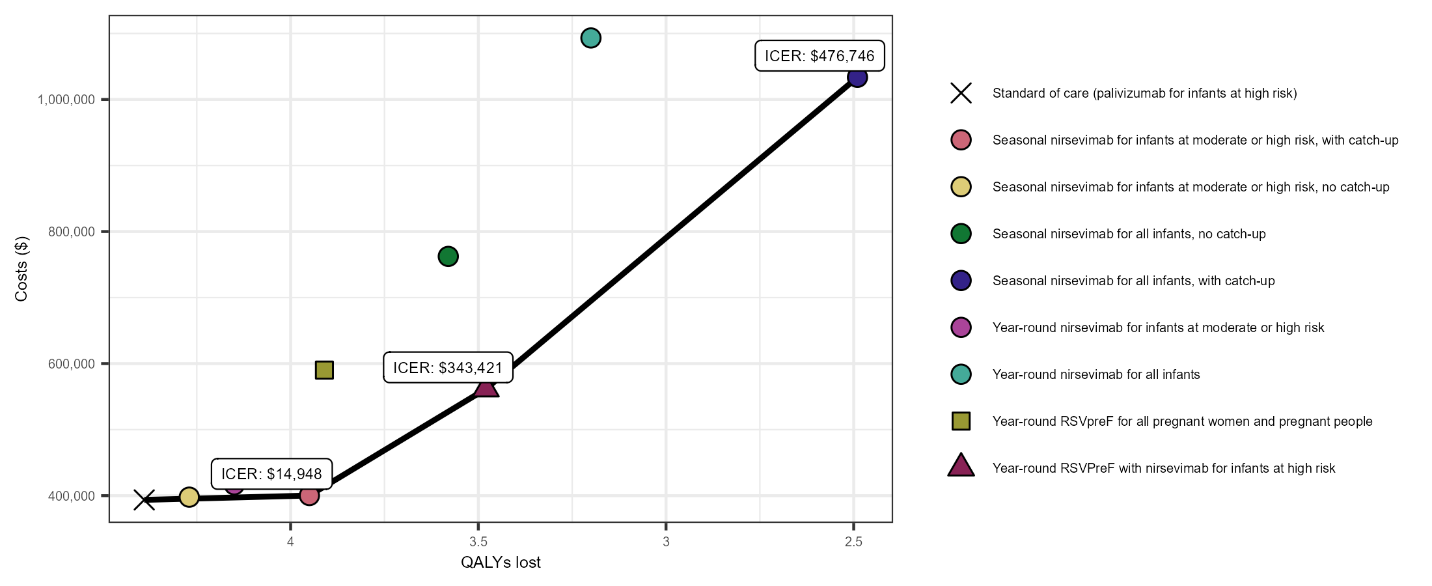

**Supplementary Figure 2:** Base case results when comparing all RSV disease prevention strategies sequentially against each other and standard of care from the societal perspective. Note: Seasonal nirsevimab for infants at moderate- and high-risk with no catch-up was dominated, but the dot representing this program appears to be on the efficiency frontier due to the scale of the graph.

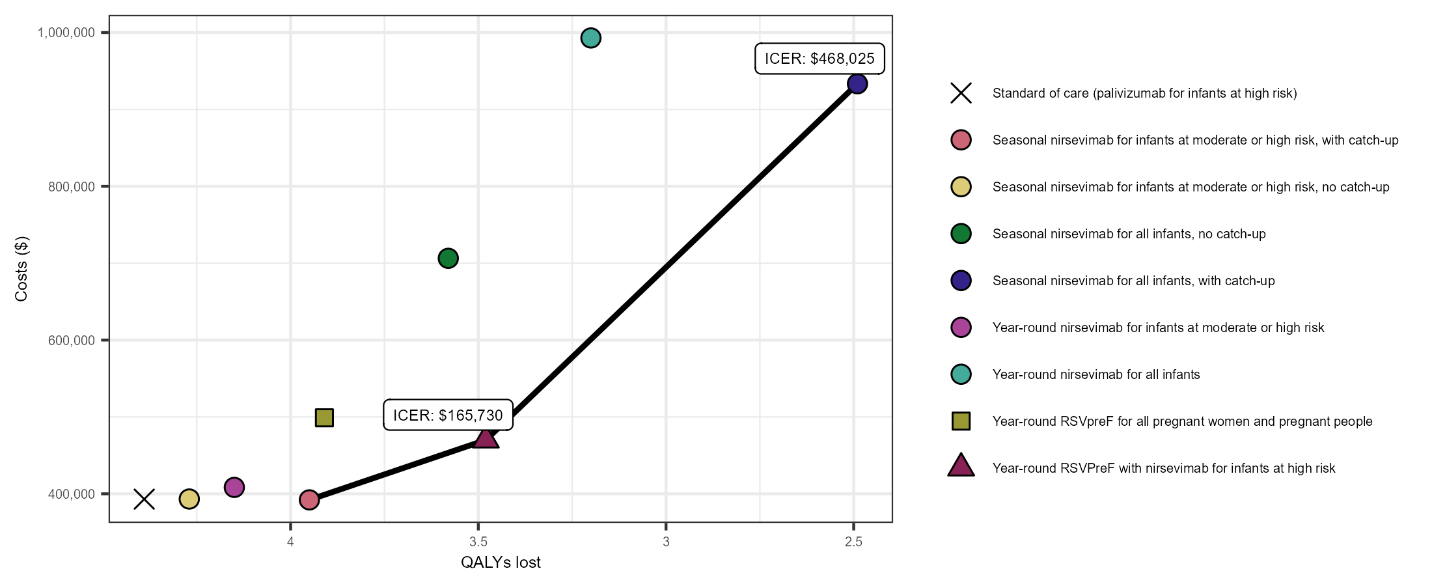

**Supplementary Figure 3:** Cost-effectiveness frontiers for no caregiver costs during immunization scenario when comparing all strategies sequentially against each other and standard of care from the societal perspective.
